# Men’s perceptions of long-term health outcomes following fertility problems: A United Kingdom-based mixed-methods survey

**DOI:** 10.64898/2025.12.12.25342139

**Authors:** Faiza Afzal, Ling Yin Fritz Wong, Mitana Purkayastha, Yan Lu, Philippa Rees, Melissa A. Richard, Carrie L. Williams, Philip J. Lupo, Barbara Luke, Michael L. Eisenberg, Allan Pacey, Alastair G. Sutcliffe

## Abstract

**Purpose:** To explore men’s assisted reproductive technology (ART) experiences, awareness and concerns about fertility-associated health outcomes, and perceptions of using administrative health records in the United Kingdom (UK) without consent to investigate these outcomes.

**Methods:** Over a two-year period, all adult men were eligible to complete an anonymous online survey distributed via a UK-based fertility charity’s social media. Free-text responses underwent thematic analysis, and categorical responses were analysed with descriptive statistics and Fisher’s exact test.

**Results:** Among 80 participants, most were aged ≥40 (66.7%), completed university (70.2%), White (77.2%) and UK residents (83.0%). Older (*p*=0.004) and White (*p*=0.001) men more likely underwent ART. Most ART users received treatment privately (60%) within the past three years (71.4%). Only one-fifth of the 15 men with identified fertility problems received discussion on fertility-associated health outcomes in clinic. Regarding perceptions, most were unaware of but concerned about these outcomes across biopsychosocial aspects, with participant quotes reflecting uncertainty and vulnerability. Recency of ART was associated with awareness (*p*=0.015) and concerns (*p*=0.001). Overall, 90.3% supported using administrative health records to investigate long-term health of fertility-challenged men, and 84.2% had no concerns about doing so without individual consent under established legal frameworks. Others raised concerns about the reliability of data anonymisation. Quotes suggested participants’ desire to understand the wider health implications of male fertility amidst a perceived gender imbalance in fertility research.

**Conclusions:** Gaps in participant knowledge, clinician communication and research in male fertility-associated outcomes support the need for universal education and further investigations in these outcomes.

**Capsule Summary:** Most men showed limited awareness of but notable concern about fertility-related long-term health outcomes, and most supported using administrative health records without individual consent for investigating these outcomes.

## Introduction

Assisted reproductive technology (ART) encompasses all treatments and procedures involving the *in vitro* handling of human oocytes, sperm or embryos with the purpose of establishing a pregnancy [1]. This includes *in vitro* fertilisation (IVF), intra-cytoplasmic sperm injection, related micromanipulation methods, and intrauterine insemination under specific circumstances [1]. An estimated 9.8–13.0 million infants have been born worldwide via ART since the first birth in 1978, with numbers increasing annually [2], and over 300,000 of these births are in the United Kingdom (UK) [3].

Male subfertility, meaning any form of reduced male fertility associated with prolonged time to conception, contributes to 30–50% of infertility cases in the UK [4, 5]. The often-overlooked scale of male contribution to infertility have been highlighted in contemporary reviews of male reproductive health [6]. While ART increases the likelihood of achieving pregnancy in affected individuals, little is known about the broader health implications of male subfertility itself. Increasing evidence suggests that male subfertility may be an early marker of underlying health conditions, including testicular cancer [7], reproductive conditions [8], diabetes [9], cardiovascular disease [10], infections [11] and mental health issues [12]. Recent concerns about the possibility of declining semen quality and fertility may reflect underlying systemic disorders [6]. These associations prompt investigation into the long-term health outcomes of subfertile men, an area that has historically received limited attention. A recent landmark review has highlighted the high prevalence and morbidity associated with male fertility issues and advocated for greater equity in research, given the current disproportionate focus on female reproductive health [13]. Equally, female infertility has been associated with long-term general health conditions [14].

To date, most research in ART has focused on perinatal and child health outcomes, with an emphasis on the safety and effectiveness of the procedures for the resulting offspring [15]. Fewer studies have investigated the health implications for men undergoing fertility treatment or the psychosocial aspects of their experience [16]. There is growing recognition that men are under-engaged in fertility care and their psychological, social and informational needs during fertility treatment are poorly understood and rarely addressed [17]. Even less is known about how healthcare professionals communicate potential health risks to men in fertility clinics, or how men perceive such information, especially in relation to emerging research on long-term health risks.

Understanding men’s perspectives is crucial, both for tailoring clinical risk communication and ensuring ethical recruitment into future studies, particularly those involving national health data. National data linkage studies, for example, offer unique opportunities to investigate associations between male subfertility and long-term health outcomes, yet little is known about how such studies are perceived by patients. This is important to establish through active patient and public involvement (PPI), as gaining individual consent for large-scale data linkage studies, particularly those utilising administrative data, may not be practical. Such studies require Section 251 support under the National Health Service (NHS) Act 2006 in England and Wales to lawfully access NHS health records for research without patient consent [18]. Our group proposes to conduct a UK register-based population cohort study to investigate the long-term general health outcomes of subfertile men, through seeking Section 251 support to access administrative health records of approximately 500,000 men without individual consent.

Therefore, this survey aims to explore men’s: (i) experience with ART if applicable, (ii) awareness of and concerns about potential adverse health outcomes associated with fertility problems and (iii) perceptions of using administrative health records without consent to investigate these outcomes.

## Methods

### Survey distribution and design

All adult men, regardless of fertility and ART status, were eligible and voluntarily completed the cross-sectional, English, anonymous survey. This survey was developed using Qualtrics and distributed through Fertility Network UK’s social media platforms (Instagram, Facebook and Twitter/X); no paid promotions were used to boost participation. The charity supports individuals facing fertility challenges across the UK, with its social media reaching more than two million users annually [19]. Additionally, the survey link was pinned on the website of our proposed population cohort study to reach a broader audience.

Before displaying the survey questions, background information was provided on the survey landing page. This outlined the survey’s rationale, objectives and target audience in non-technical language. We broadly described our proposed study’s intended methodology including Section 251 support, without mentioning the study name, to elicit perceptions more generally and avoid bias [20]. A disclaimer was included to inform participants that some questions might address sensitive areas, with signposting to the Fertility Network UK helpline for those affected by the survey content. Informed consent was obtained from all participants prior to their inclusion in the survey. This study was granted approval by the ethics committee of the authors’ primary institution (reference number: 22/LO/0536). The survey did not collect any personal identifiable data and remained open for two years from 18 August 2022 to 14 July 2024.

The survey was structured into four domains to align with our objectives (Table 1). Domain 1 collected demographic details. Domain 2 studied respondents’ experience with fertility treatment. Domain 3 investigated their concerns related to potential adverse health outcomes. Domain 4 explored their perceptions of research methodology. Questions were a combination of open-ended questions (Q5, 7, 8 and 18) and multiple-choice questions (Q1–4, 6 and 9–17). All survey questions were optional given the sensitive nature of the topic.

**Table 1.** Domains and item response rates of survey questions.

| Domain | Survey question | Actual responses/<br>expected responses<br>(item response rate) |
| --- | --- | --- |
| Domain 1:<br>Demographics | Q13: How old are you? | 57/80 (71.3%) |
|  | Q14: What is your highest level of education? | 57/80 (71.3%) |
|  | Q15: What is your ethnic group? Choose one option that best describes your ethnic group or background. | 57/80 (71.3%) |
|  | Q16: Are you a UK resident? | 47/80 (58.8%) |
| Domain 2:<br>Experience<br>with fertility<br>treatment | Q1: Are you considering assisted reproduction treatment, or have you undergone assisted reproduction treatment to conceive a child (whether or not this is or was due to male fertility problems)? | 79/80 (98.8%) |
|  | Q2: How long ago was it? If you have had more than one treatment, tell us about your last treatment. | 35/35 (100.0%) |
|  | Q3: Was it at a Private or NHS clinic? By this we mean the type of clinic rather than the type of funding for the treatment(s). | 34/35 (97.1%) |
|  | Q9: Thinking again about your fertility treatment, was fertility evaluation (e.g. semen analysis, hormone or genetic testing, physical examination) performed at this clinic before the treatment? | 27/35 (77.1%) |
|  | Q10: Were the results of this fertility evaluation discussed with you at this clinic? | 22/22 (100.0%) |
|  | Q11: Did the results of this fertility evaluation indicate that you may have fertility problems? | 19/19 (100.0%) |
|  | Q12: Were potential adverse health outcomes associated with fertility problems discussed with you at this clinic? | 15/15 (100.0%) |
| Domain 3:<br>Potential<br>adverse health<br>outcomes in<br>men<br>associated<br>with fertility<br>problems | Q4: Are you aware of potential adverse health outcomes in men associated with fertility problems? | 76/80 (95.0%) |
|  | Q5: To the best of your knowledge, what medical conditions or illnesses are associated with potential fertility problems in men? | 15/22 (68.2%) |
|  | Q6: Are you concerned about potential adverse health outcomes in men associated with fertility problems? | 70/80 (87.5%) |
|  | Q7: What are your concerns about potential adverse health outcomes in men associated with fertility problems? | 39/49 (79.6%) |
| Domain 4:<br>Perceptions of<br>research<br>methodology | Q8: As described, our research team will use detailed administrative health records to study the long-term health of men in the UK affected by fertility problems, what are your views on the benefits of such a study? | 62/80 (77.5%) |
|  | Q17: As described, we are seeking support by Section 251 of the National Health Service Act 2006 to access information about participants in this study (around 500,000 men) without consent. Section 251 of the NHS Act 2006 allows the use of confidential patient information for medical research when it is not possible to use anonymised information or when seeking consent is not practical. Do you have any concerns about using confidential patient information without consent for the purpose of this study? Please note that at no point will the research team have access to information that allows men | 38/80 (47.5%) |
|  | included in this study to be identified. |  |
|  | Q18: What are your concerns about using confidential patient information without consent for the purpose of this study? Is there anything we can do to reassure you about this? | 5/6 (83.3%) |

Some follow-up questions were conditional and only displayed if respondents answered 'yes' to a preceding question (skip logic). The level of indentation in the table corresponds to the level of hierarchy within the questions. The number of expected responses reflects this skip logic when calculating the item response rates.
Abbreviations: HFEA, Human Fertilisation and Embryology Authority; NHS, National Health Service; Q, question number; UK, United Kingdom.

### Data analysis

Quantitative analysis of categorical responses (Q1–4, 6 and 9–17) was performed to produce descriptive statistics. Due to small sample sizes and empty cells, Fisher’s exact test was conducted to compare group differences, with statistical significance set at two-tailed *p* < 0.05. Sensitivity analysis including missing responses was performed to assess the impact of survey completion on the results. All analyses were performed on StataNow/MP 18.5.

The remaining free-text responses (Q5, 7, 8 and 18) were analysed qualitatively using an inductive and semantic approach to thematic analysis [21], where themes were derived from the data and reflected the explicit meaning of responses. Themes were then triangulated with quantitative findings under a post-positivist framework. To enhance the credibility of the analysis, we followed Nowell et al.’s guidelines to establish trustworthiness [22]. The second author manually coded all responses in NVivo 14 and organised the codes into themes, with reflexive memos and audit trails documented throughout. The resulting codebook was independently reviewed by the first and last authors. Code frequencies were counted and converted to percentages to enable comparison across questions. Illustrative quotes were included to support the findings.

## Results

### Domain 1: Demographics

Among the 109 responses submitted, 29 submissions provided empty answers to all questions and were excluded. This resulted in 80 responses for analysis (completion rate 73.4%), though item response rates varied across individual questions (Table 1).

Across all ART statuses, most respondents were aged 40 or above (66.7%), held a university degree or higher (70.2%), were White (77.2%) and a UK resident (83.0%) (Table 2). A relatively even distribution of ART status was captured, with 44.3% having undergone ART. All men who had undergone ART were White and completed at least high school or college. Significant differences were observed in men’s ART status by age (*p*=0.004) and ethnicity (*p*=0.001): older and White men were more likely to have undergone ART. Sensitivity analyses including missing responses showed no change in significance.

**Table 2.**
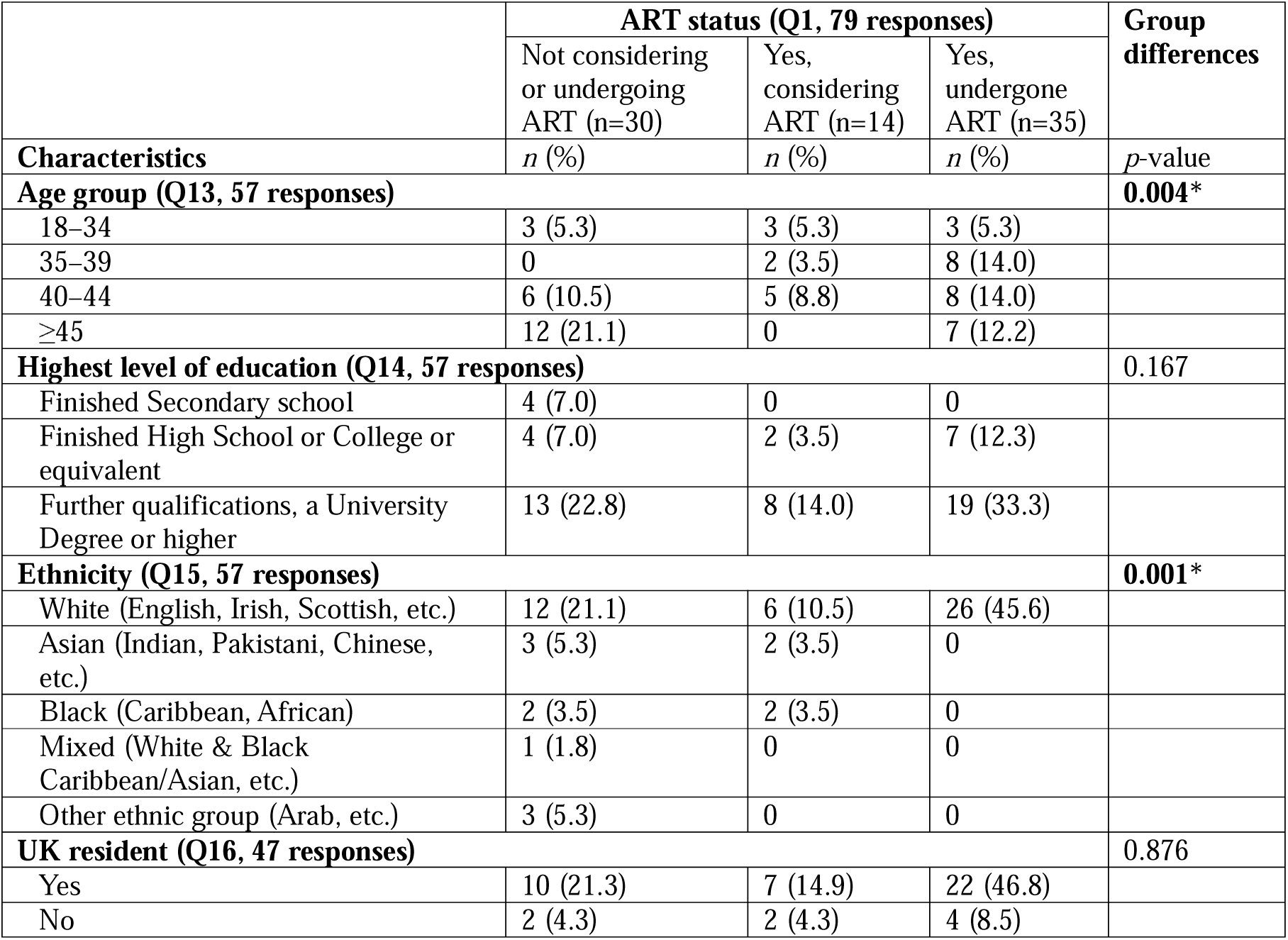
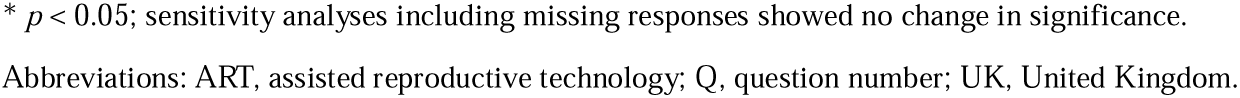
Respondent characteristics (n=79).

|  | ART status (Q1, 79 responses) |  |  | Group differences |
| --- | --- | --- | --- | --- |
|  | Not considering or undergoing ART (n=30) | Yes, considering ART (n=14) | Yes, undergone ART (n=35) |  |
| Characteristics | n (%) | n (%) | n (%) | p-value |
| **Age group (Q13, 57 responses)** |  |  |  | **0.004\*** |
| 18–34 | 3 (5.3) | 3 (5.3) | 3 (5.3) |  |
| 35–39 | 0 | 2 (3.5) | 8 (14.0) |  |
| 40–44 | 6 (10.5) | 5 (8.8) | 8 (14.0) |  |
| ≥45 | 12 (21.1) | 0 | 7 (12.2) |  |
| **Highest level of education (Q14, 57 responses)** |  |  |  | 0.167 |
| Finished Secondary school | 4 (7.0) | 0 | 0 |  |
| Finished High School or College or equivalent | 4 (7.0) | 2 (3.5) | 7 (12.3) |  |
| Further qualifications, a University Degree or higher | 13 (22.8) | 8 (14.0) | 19 (33.3) |  |
| **Ethnicity (Q15, 57 responses)** |  |  |  | **0.001\*** |
| White (English, Irish, Scottish, etc.) | 12 (21.1) | 6 (10.5) | 26 (45.6) |  |
| Asian (Indian, Pakistani, Chinese, etc.) | 3 (5.3) | 2 (3.5) | 0 |  |
| Black (Caribbean, African) | 2 (3.5) | 2 (3.5) | 0 |  |
| Mixed (White & Black Caribbean/Asian, etc.) | 1 (1.8) | 0 | 0 |  |
| Other ethnic group (Arab, etc.) | 3 (5.3) | 0 | 0 |  |
| **UK resident (Q16, 47 responses)** |  |  |  | 0.876 |
| Yes | 10 (21.3) | 7 (14.9) | 22 (46.8) |  |
| No | 2 (4.3) | 2 (4.3) | 4 (8.5) |  |

\* $p < 0.05$ ; sensitivity analyses including missing responses showed no change in significance.
Abbreviations: ART, assisted reproductive technology; Q, question number; UK, United Kingdom.

### Domain 2: Experience with fertility treatment

Further insights were gathered from follow-up questions directed at the subset of 35 respondents who had undergone ART.

### ART clinic and timeframe

More than half (n=21, 60%) attended a private clinic (Table 3). The timing of treatment varied, with most having received treatment within the past three years (n=25, 71.4%). Two reported receiving treatment 3–5 years ago, both completed privately, while eight had treatment over five years ago.

**Table 3.** ART clinic type and timeframe of respondents who had undergone ART (n=35).

|  | ART clinic type (Q3, 34 responses) |  |  |
| --- | --- | --- | --- |
| | NHS clinic<br>($n=13$) | Private clinic<br>($n=21$) | Did not answer<br>($n=1$) |
| ART timeframe (Q2, 35 responses) | $n$ (%) | $n$ (%) | $n$ (%) |
| <1 year ago | 6 (17.1) | 10 (28.6) | 0 |
| 1–3 years ago | 4 (11.4) | 5 (14.3) | 0 |
| 3–5 years ago | 0 | 2 (5.7) | 0 |
| >5 years ago | 3 (8.6) | 4 (11.4) | 1 (2.9) |

Abbreviations: ART, assisted reproductive technology; Q, question number.

### Fertility evaluation

Prior to treatment, most men (n=22, 81.5%) underwent a fertility evaluation, and results were discussed with most of them (n=19, 86.4%) (Fig. 1). Of these, 15 (78.9%) men were informed of fertility problems, while four were not. However, among those with identified fertility problems, only one-fifth (n=3) reported that potential adverse health outcomes were discussed at the fertility clinic, while the remaining reported no such discussion.

**Fig. 1.**
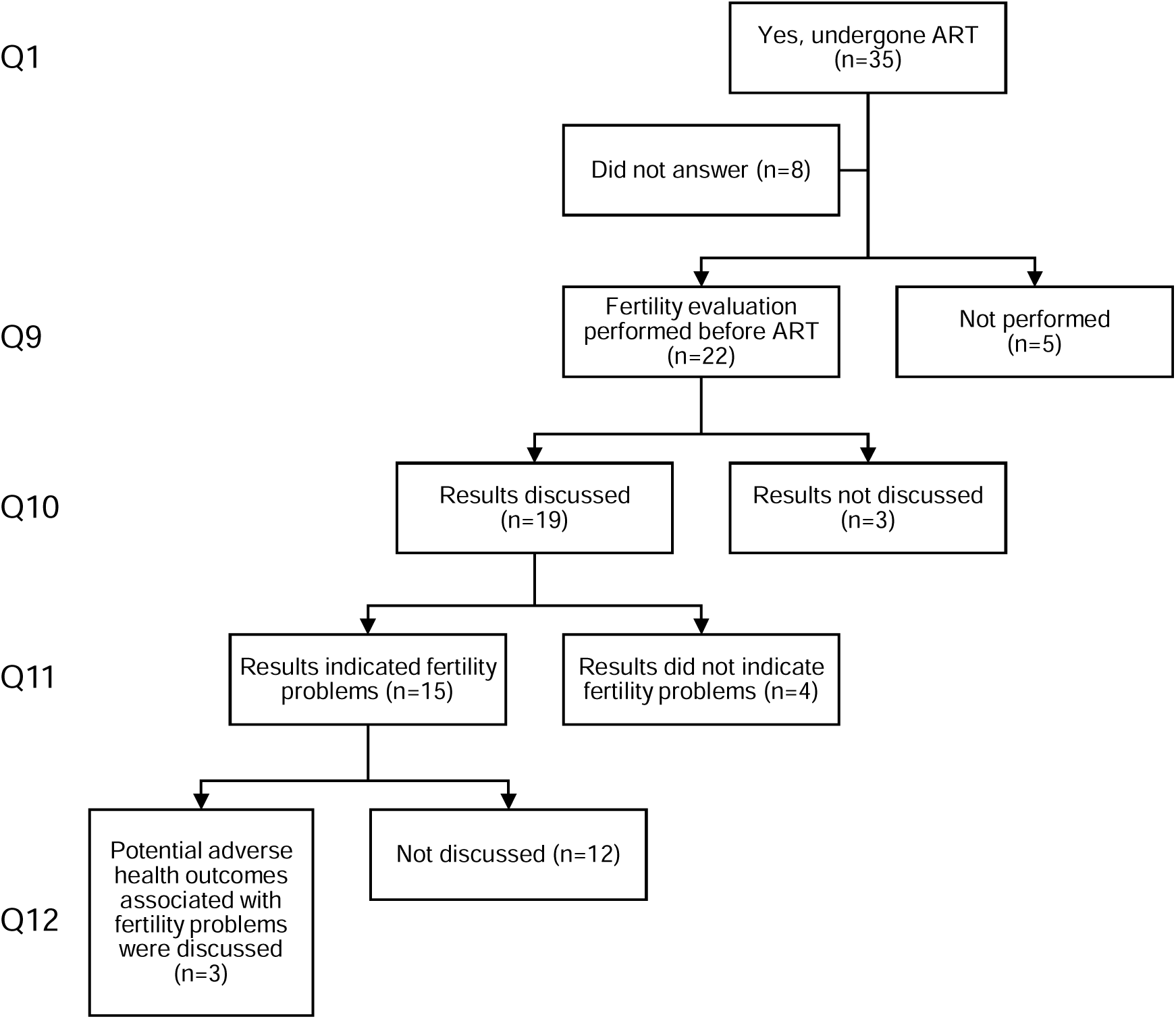
Flowchart of fertility evaluations and related discussions among men who had undergone ART (n=35). Responses detail whether fertility evaluation was performed, whether results were discussed and whether potential adverse health outcomes associated with fertility problems were addressed. Each level down the hierarchy represents a conditional follow-up question to the one preceding it. Abbreviations: ART, assisted reproductive technology; Q, question number.

### Domain 3: Most men were unaware of but concerned about fertility-related health outcomes in biopsychosocial aspects

Nearly half of the respondents (34 out of 69 responses in both Q4 and Q6, 49.3%) were unaware of, but were concerned about, potential adverse health outcomes in men associated with fertility problems. Fifteen men (21.7%) were both concerned and aware, and 18 (26.1%) were neither concerned nor aware. However, there was no association between having awareness and concerns (*p*=0.122). Respondent characteristics did not differ between those who answered and did not answer questions in this domain (all *p*>0.05).

Among the various characteristics examined in Supplementary Tables 1–2, the recency of ART was significantly associated with awareness (*p*=0.015) and concerns (*p*=0.001). All men who reported awareness had undergone ART within the past year. Similarly, all men treated within the past year expressed concerns, with proportions declining as time since treatment increased.

### Awareness

Most men (54 out of 76 responses, 71.1%) were unaware of potential adverse health outcomes in men associated with fertility problems. The remaining participants identified 25 conditions or illnesses, with a predominance of physical over mental health issues. Awareness was significantly more common among non-UK residents than UK residents (*p*=0.018) (Supplementary Table 1), although the categories of issues identified did not differ between the two groups (*p*=0.271).

Reproductive issues were most frequently mentioned (Fig. 2), including a variety of conditions, particularly low sperm quality (n=6), DNA fragmentation (n=3) and varicocele (n=2). Other reproductive issues were mentioned only once. Participant responses reflected a nuanced understanding, including the use of specific terminology such as *‘oligozoospermia’* (P53, age 40–44, undergone ART) and distinctions between *‘sperm morphology and motility’* (P44, age 40–44, considering ART). Cancer and cardiovascular conditions were next most common (each n=7), followed by infection (n=5) and mental health issues (n=4).

**Fig. 2.**
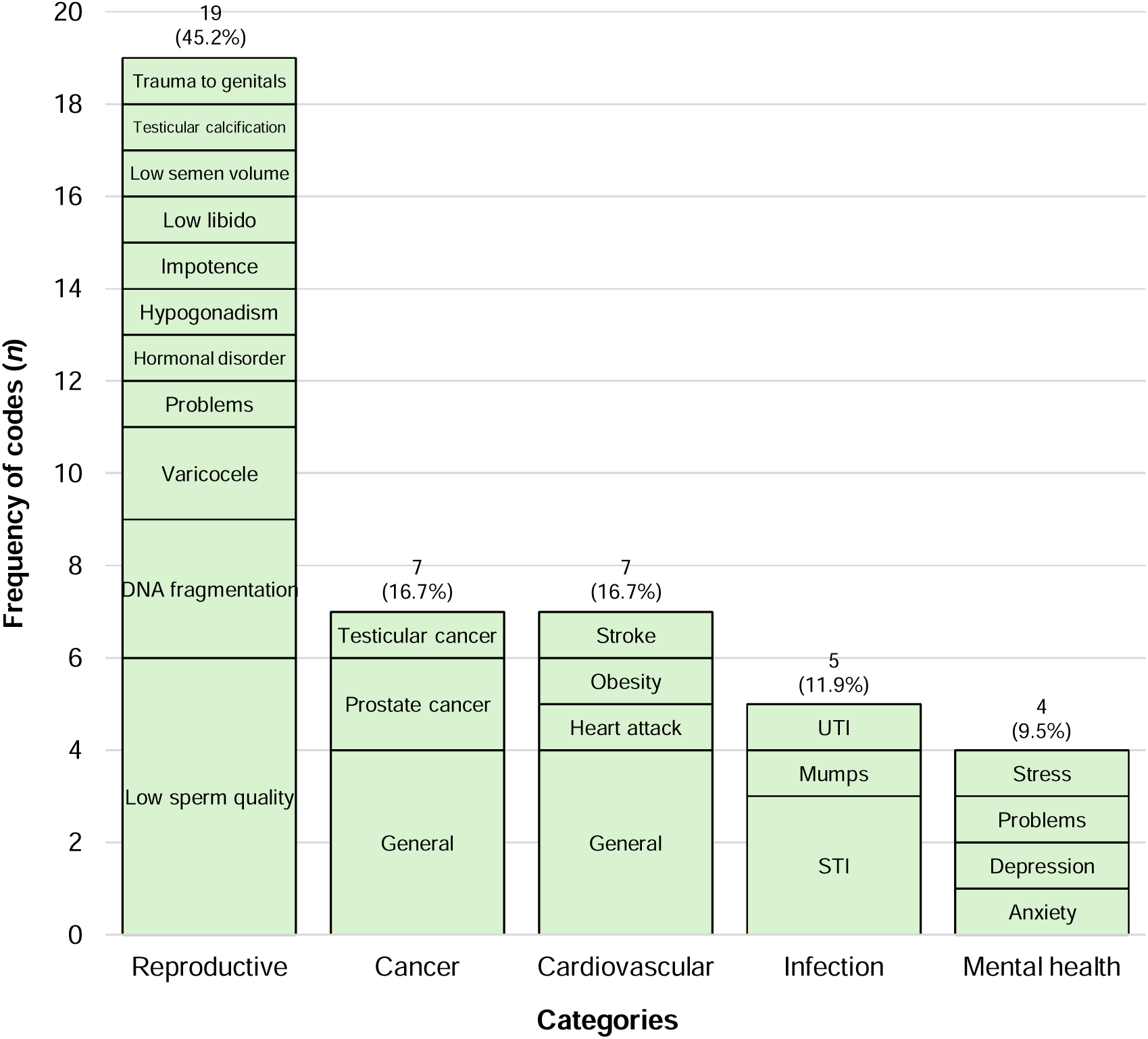
Men’s knowledge of medical conditions associated with potential fertility problems in men (Q5). Labels within bars represent codes generated from thematic analysis, and the labels above bars represent the total frequency and percentage of codes for each category. ‘General’ indicates responses that referred broadly to a category without identifying a specific condition. Abbreviations: STI, sexually transmitted infections; UTI, urinary tract infection.

### Concerns

Most men (49 out of 70 responses, 70%) reported concerns about potential adverse health outcomes associated with fertility problems. These concerns covered physical and mental health, and often reflected feelings of uncertainty and vulnerability. Ethnicity was significantly associated with the expression of concern (*p*=0.007), with 80% of White men and all Black men reporting concern (Supplementary Table 2).

Most themes related to general health (n=15, 30.6%) (Fig. 3), particularly long-term outcomes and whether fertility issues indicated part of a larger problem: *‘That they could limit lifespan and therefore the amount of time available to spend with one’s children’* (P56, age 40–44, undergone ART), *‘What impact this could have on the rest of my life outside of the fertility problems’* (P18, age 18–34, undergone ART). Reproductive concerns followed (n=10, 20.4%), including low sperm count, low testosterone and offspring health, as one participant noted: *‘if lucky to have a child [and] if the child will inherit any of these health outcomes’* (P53, age 40–44, undergone ART). Among mental health (n=10, 20.4%) concerns, men described the emotional toll of fertility problems particularly on relationships, for example, as *‘a strain on families, marriages and partnerships’* (P28, age 18–34, considering ART). Another respondent reflected on the implications of age in fatherhood, noting that *‘parenting at an older age may be more stressful and may have less social support available’* (P19, age 18–34, not considering or undergoing ART). Equally, not knowing (n=7, 14.3%) itself was also a source of concern, with the frustration of not knowing and not told expressed: *‘That I don’t know what they are. This has never been communicated to me’* (P72, age 35–39, undergone ART). Notably, although cardiovascular disease, cancer and infection were acknowledged as potential outcomes in Q5, fewer men cited them as concerns in Q7.

**Fig. 3.**
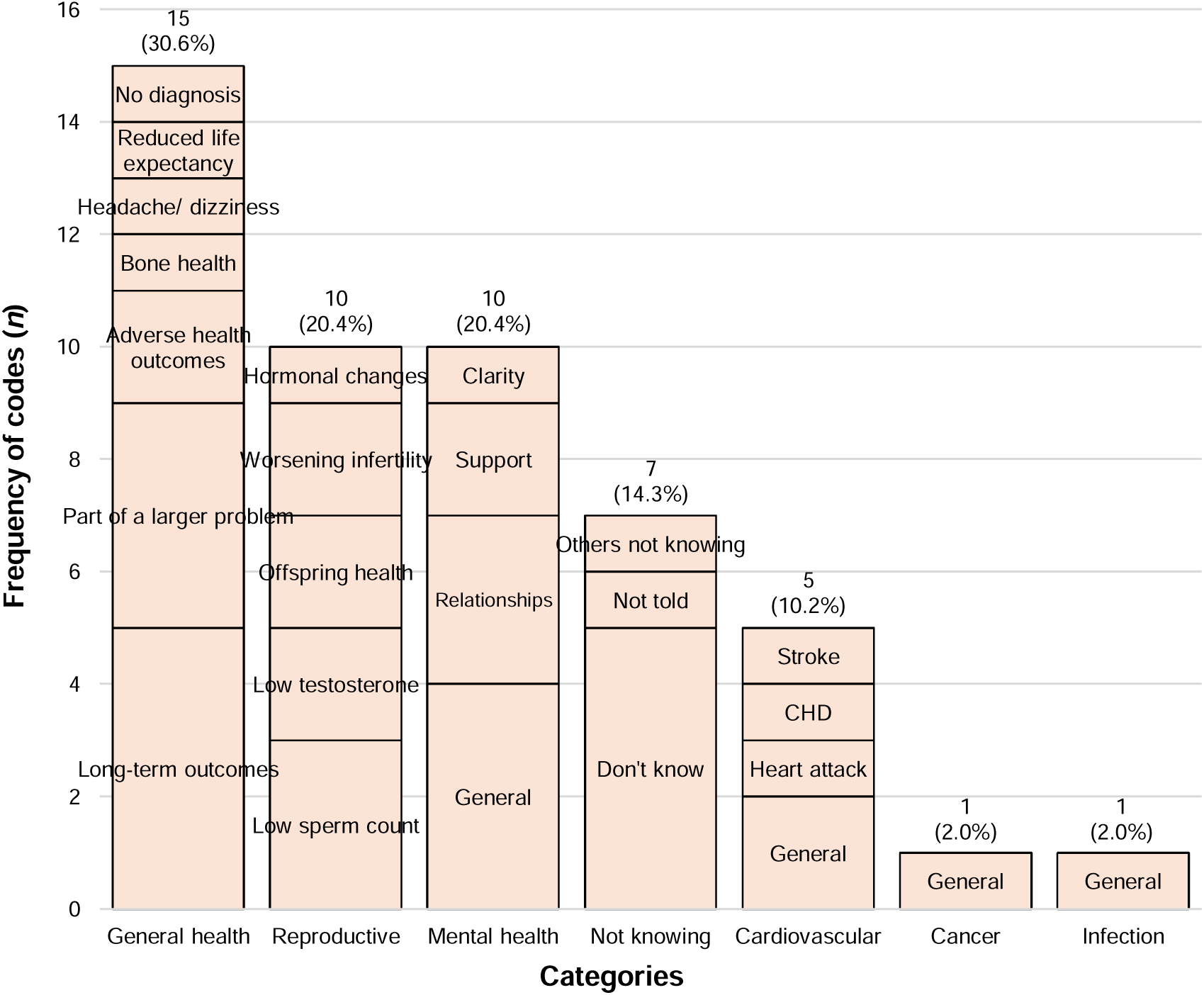
Men’s concerns about potential adverse health outcomes associated with fertility problems (Q7). Labels within bars represent codes generated from thematic analysis, and the labels above bars represent the total frequency and percentage of codes for each category. ‘General’ indicates responses that referred broadly to a category without identifying a specific condition. Abbreviation: CHD, coronary heart disease.

### Domain 4: Overall positively perceived benefits of using administrative health records without consent for studying long-term health of fertility-challenged men

Participant perceptions of the benefits of using administrative health records to study the long-term health of fertility-challenged men were broadly categorised as positive (n=56, 90.3%), conditional (n=4, 6.5%) and negative (n=2, 3.2%) (Table 4). Among the positive perceptions, most participants expressed general support (25.8%), though typically provided without additional elaboration or detail. This was followed by more specific themes on the benefits of research to patients and the field: to generate more information (17.7%), address the research gap in male infertility (11.3%) and enable treatment options (9.7%). Others felt the study could raise awareness (8.1%) and highlight the potential long-term health outcomes of fertility problems (6.5%). Overall, participant quotes reflected altruism, optimism and a desire to better understand male fertility and its broader health implications: *‘There’s never been an “answer” as to why I’m infertile and our knowledge of male fertility seems so limited … I’d be interested to know if there’s any risk of prostate issues, cancer, anything else due to infertility – maybe I’ll live longer – so pleased you’re carrying this project out’* (P50, age ≥45, undergone ART). Some quotes reflected a perceived gender imbalance in fertility research and public awareness, with men’s health often seen as secondary (P3, P43 and P78 in Table 4). Four respondents expressed conditional support which depends on anonymity and informed participation. Negative views were uncommon, with only two participants citing concerns about confounding and relationship strain.

**Table 4.** Participant perceptions of using administrative health records to study the long-term health of men in the UK affected by fertility problems (Q8).

| Perception/code | Frequency (%) | Illustrative quote |
| --- | --- | --- |
| **Positive perceptions** |  |  |
| General support | 16 (25.8) | *'Anything that adds onto a subject like this will be a good thing.'* (P63, age 40–44, considering ART) |
| Generates more information | 11 (17.7) | *'To show any previously unidentified issues linked with fertility issues.'* (P12, age $\geq 45$, not considering or undergoing ART) |
| Addresses research gap in men infertility | 7 (11.3) | *'I guess similar research is focused mostly on women and we know little about men.'* (P3, age 40–44, not considering or undergoing ART) |
| Enables treatment options | 6 (9.7) | *'This research would also be beneficial in proffering possible intervention treatments to the affected men.'* (P28, age 18–34, considering ART) |
| Raises awareness | 5 (8.1) | *'Potentially putting male health in the headlines. It seems men's health is a side issue currently.'* (P43, age 35–39, considering ART) |
| Highlights long-term outcomes | 4 (6.5) | *'Not all men want to have kids who have fertility issues. We need to still be aware of long-term issues even if that's the case for future health.'* (P59, age $\geq 45$, not considering or undergoing ART) |
| Enables early detection | 3 (4.8) | *'Could lead to early detection of serious previously unknown conditions.'* (P14, age $\geq 45$, not considering or undergoing ART) |
| Enables future research | 2 (3.2) | *'Useful for future medical advances.'* (P70, age 18–34, undergone ART) |
| Addresses stigma | 1 (1.6) | *'This study should also help address the stigma surrounding fertility issues.'* (P19, age 18–34, not considering or undergoing ART) |
| Enables emotional support | 1 (1.6) | *'It can help woman to understand more and to support more their men in their fertility journey. It can help men to understand themselves and their challenges better.'* (P78, age 35–39, undergone ART) |
| **Conditional perceptions** |  |  |
| If anonymity is maintained | 3 (4.8) | *'Beneficial to society as long as individual privacy is maintained.'* (P58, age 18–34, considering ART) |
| If participants are informed | 1 (1.6) | *'This will indeed help collect valuable information but how will the men whose data has been collected be informed?'* (P34, age $\geq 45$, not considering or undergoing ART) |
| **Negative perceptions** |  |  |
| Adds tension to relationships | 1 (1.6) | *'It would give more information towards a decision on treatment. This may deter males who would have agreed to be more cautious, adding to the tension partners feel in the process of accepting infertility and seeking assistance.'* (P30, age $\geq 45$, undergone ART) |
| Concerns over confounders | 1 (1.6) | *'These studies are always hard to do as there will be many confounding factors involved.'* (P18, age 18–34, undergone ART) |

Abbreviations: ART, assisted reproductive technology; P, participant number.

Moreover, most respondents (n=32, 84.2%) had no concerns about using confidential patient information without consent for the purpose above, via Section 251 support where required. Concerned individuals (n=6, 15.8%) raised issues around data anonymisation and the need for explicit consent. Participants voiced scepticism around the reliability of anonymisation: *‘Working in IT Security, and knowing of previous data breaches in civil servant departments, I don’t trust that data will be properly anonymised and not accidently breached.’* (P49, age ≥45, undergone ART). Others felt that using personal information without consent would be *‘a huge invasion of privacy into an incredibly emotionally difficult subject matter’* (P65, age 40–44, not considering or undergoing ART), while one suggested that *‘even brief consent by post would be preferable’* (P80, age 35–39, considering ART).

## Discussion

Men’s perceptions of fertility-related health outcomes and research methodologies for studying these long-term outcomes were explored in the context of their fertility experiences. Our findings show that: (i) men were unaware of but concerned about various fertility-related health outcomes; (ii) such awareness and concerns were associated with how recently they had ART and other factors; and (iii) the majority supported using administrative health records and confidential patient data without consent for research on long-term outcomes in men affected by fertility problems. These findings offer novel insights into men’s fertility-related health perceptions and expectations of fertility research.

Nearly three-quarters of respondents were unaware of fertility-related health outcomes, highlighting a significant gap in male fertility knowledge. A population-based survey of Canadian men [23] similarly found that they could only identify less than half of the health issues associated with male infertility. Consistent with their findings and contrary to expectations, non-UK residents showed greater levels of knowledge while other demographic differences in awareness were small [23], potentially reflecting variations in health literacy or cultural attitudes. From a broader perspective, research participants are typically known to be better educated than the general population (‘healthy volunteer bias’) [24], yet our respondents largely remained unaware of potential fertility-related risks. While generalisation is statistically limited with our sample size, this picture may suggest that the awareness in the general male population may be even lower, further hindering their health-seeking behaviours and participation in reproductive services [25]. Nonetheless, those who were aware demonstrated knowledge broadly consistent with the established risks. Respondents identified reproductive conditions [8], male-specific cancers [26], cardiovascular disease [10], infections [11] and mental health issues [12]. These associations have been supported by systematic reviews and align with the current understanding that impaired male reproductive function may serve as a proxy for poor general health [27–29]. Kasman et al. [30] further elucidate the bidirectional relationship of these associations with male infertility. It is, therefore, ever more important to raise awareness among men regarding these comorbidities. The European Association of Urology [31] and American Urological Association/American Society for Reproductive Medicine [32] guidelines recommend that ‘clinicians should counsel infertile men or men with abnormal semen parameters of the health risks associated with abnormal sperm production’, yet in our survey, only one-fifth of men with identified fertility problems received such discussions. This may reflect a gap in the UK National Institute for Health and Care Excellence guidelines [33], which make no mention of such counselling for men. All respondents who reported awareness had undergone ART within the past year, suggesting that awareness may diminish over time and such counselling may be most effective when delivered around the time of treatment. Regular, updated discussions are essential to sustain long-term awareness. As the low awareness was observed across most demographic groups, our results also call for universal education on fertility-related adverse health outcomes in men as per previous studies [23].

Regarding concerns, most men were concerned about the impact of fertility problems on their physical and mental health. Such expression of concern was associated with ethnicity, consistent with prior surveys [34, 35]. However, this association may be mediated by factors not captured in our study, considering the broader evidence confirming the ethnic or geographical differences in semen parameters [36], male infertility prevalence [37], consanguinity rates [38] and cultural stigma towards infertility [39]. Because infertility is multifactorial [40], it is difficult to discern the specific reasons behind the ethnic differences in concern observed here. Overall, unlike the more concealed [41, 42] or short-term [43] concerns reported by men in other studies, our respondents readily voiced concerns which were mainly biological, focusing on long-term general and reproductive health outcomes. This may be attributable to their recent engagement with ART [44], different study designs [12] or personal experiences [41]. Although the literature on men’s specific biological concerns is scarce, their pattern of concerns mirrored the conditions men reported being aware of, suggesting that awareness and concerns tend to focus on the same domains. Psychosocially, men’s concerns about relationships [34, 45], support [46] and uncertainty of not knowing [47] have been well-established and associated with increased anxiety and depression. These psychological effects have, in turn, been linked to poorer sperm quality [48–50], greater partner stress [47] and reduced help-seeking [51], potentially creating a vicious cycle that intensifies men’s distress surrounding fertility-related adverse outcomes. These emotions occur against a background of men’s desire for fatherhood [52] and need for information as they cope with uncertainty while remaining hopeful [53], as reflected in participant quotes. The discordance between low awareness and high concern rates in our survey indicates a clear unmet need for information, emotional support and improved clinician communication across the ART pathway, echoing the findings of Winston et al.’s systematic review [54]. Clinicians should actively listen to and address men’s most common concerns, not only from a medical but also psychosocial perspective [54].

Apart from clinical implications, our findings highlight research implications for using administrative health records and confidential patient data without consent for studying long-term outcomes in men affected by fertility problems. Firstly, regarding data usage, respondents’ primarily positive perceptions, despite some concerns over privacy and consent, reflect a similar proportion of the public attitudes across 20 UK-based general-setting studies included in Stockdale et al.’s systematic review [55]. In the fertility setting, however, literature on such perceptions remains limited. Among the 54 studies included across both Hutching et al.’s systematic reviews focusing on consent [56] and privacy/trust [57] in the public’s attitudes towards data usage, only one study [58] was conducted in the fertility context. This gap means that patient and public perceptions on data usage for fertility research may remain poorly characterised. Secondly, regarding data usage for male-specific fertility research, respondents supported its benefits of generating information on comorbidities [59], long-term post-ART outcomes for men [60] and their children [13, 61], and benefitting treatment and prevention strategies [62], reinforcing current research priorities in male fertility. These priorities, however, also connect to a broader issue of gender imbalance in fertility research as highlighted by respondents: even studies aimed at identifying research priorities in infertility and ART have included only 10–20% men in their samples [60, 63], indicating an underrepresentation of male perspectives. Considering both gaps, there is a clear need to investigate patient perceptions of data usage not only in fertility research, but specifically, male fertility research. Active PPI of men can help co-design research priorities and methodology that reflect their needs (‘nothing about us without us’) [64]. Our study contributes to this principle by reporting men’s perceptions, hoping to stimulate discussion on governance practices that could maximise research utility and inclusivity for men in the UK. On the other hand, trust (or lack thereof) in data sharing emerged as a small but noteworthy theme for respondents’ conditional support in this study and Aitken et al.’s systematic review [65]. Some supported data use on the condition that confidentiality and data anonymisation was assured, reflecting concerns about transparency and possible data breaches [65]. Strategies such as personalised control over data sharing, dynamic consent models and regular dissemination of results through PPI avenues have been recommended to establish trust [66], with scope for their application in male fertility research. Strengths of our study include its broad coverage of topics across four domains and its mixed-methods design. Free-text responses generated richer and more diverse qualitative insights than structured questions typically allow [67], helping to address a gap in ART research that often prioritises clinical success over patient experience [68]. Conducted over two years, the data included relatively recent ART experiences, potentially reducing recall bias [69]. The anonymity of online responses likely encouraged more honest reflection on sensitive or stigmatised topics such as mental health concerns, reducing social desirability bias. Recruited from social media users engaging with the charity’s channels, our sample had a balanced representation of men who had and had not considered or undergone ART, capturing a wider range of fertility experiences rather than focusing solely on treatment users [70]. Overall, this study contributes to an under-explored area by specifically focusing on men’s perspectives regarding fertility and health.

However, this study has important limitations. Regarding our sample, the size was relatively small as recruitment mainly relied on one charity, although this survey was kept active for two years and a similar ART-focused survey for women has achieved higher participation using the same charity [71]. This aligns with evidence that men are typically underrepresented in fertility-related surveys compared to women [60, 63, 72]. The predominantly White, highly educated sample may limit generalisability, though this reflects the sociodemographic distributions of fertility treatment users in the UK [73]. Voluntary participation may have introduced self-selection bias [74], with men who had more adverse experiences potentially overrepresented. Regarding survey design, the cross-sectional design cannot assess changes in perceptions during fertility treatment. As all questions were optional, missing responses were expected. Item response rates generally decrease as the survey progresses [75], so questions about demographics and research methodology perceptions had relatively low response rates, and their findings should thus be interpreted with caution. Questions about perceptions of research methodology could also have been phrased more clearly, as ambiguity around the term anonymisation may influence participants’ attitudes [65]. Additional variables, such as infertility type (male, female or mixed factor) and sexual orientation, may also have influenced participants’ perspectives, but these were not collected as they fall beyond the study’s primary scope. Similarly, concerns could have been quantified using established scales such as the Fertility Problem Inventory [76] to allow cross-comparison of findings with other studies. Regarding our findings, while free-text responses were independently reviewed by three researchers, qualitative interpretation remains inherently subjective and some nuances may have been overlooked. Because infertility is multifactorial [40] and subgroup sizes were small or sometimes zero, statistically significant associations by residency or ethnicity may not imply clinical significance. As a survey conducted in the UK, findings may not generalise to other countries where ART access and sociocultural contexts differ [77].

Future research should explore male fertility perceptions in more diverse and comparative contexts. Comparing men’s and women’s fertility-related concerns [43] could clarify gender differences in experiences. Qualitative longitudinal research using interviews or focus groups could provide deeper insights into how men’s views evolve across treatment stages. International, multicentre surveys would enable exploration of sociocultural and healthcare variations in male fertility awareness. Greater ethnic and socioeconomic diversity is also needed to better capture variations in public perceptions of fertility-related health outcomes [70]. Using translated survey materials, multiple recruitment sources and collaboration with community liaison workers could achieve broader representation of Black, Asian and Minority Ethnic (BAME) populations [78]. Finally, as PPI has been shown to shape the direction of both laboratory [79] and clinical research [80], incorporating PPI into future ART outcome studies can ensure they reflect men’s needs.

## Conclusion

This UK-based mixed-methods survey found that (i) most men were unaware of but concerned about male fertility-associated health outcomes; (ii) such awareness and concerns were associated with the recency of ART and other demographic factors; and (iii) most supported using administrative health records without consent for investigating long-term health outcomes in men with fertility problems. Together, our findings contribute to the limited evidence on men’s perspectives in ART experience and data usage in male fertility research. Identified gaps in participant knowledge, clinical communication and male fertility research strengthen the need for universal education and further investigations in male fertility-associated health outcomes.

## Acknowledgments

The authors thank all participants who responded to the survey and Fertility Network UK for facilitating its distribution.

All research at Great Ormond Street Hospital NHS Foundation Trust and UCL Great Ormond Street Institute of Child Health is made possible by the NIHR Great Ormond Street Hospital Biomedical Research Centre. The views expressed are those of the author(s) and not necessarily those of the NHS, the NIHR or the Department of Health.

## Declarations

### Ethics approval

The questionnaire and methodology for this study were approved by the University College London Research Ethics Committee (22/LO/0536).

### Consent to participate

Informed consent was obtained from all individual participants included in the study.

### Data availability statement

The data that support the findings of this study are not publicly available due to reasons of sensitivity and are available upon reasonable request from the corresponding author, F.A. Data are located in controlled access data storage at University College London.

### Author contributions

F.A. and L.Y.F.W. have contributed equally to this work and share first authorship.

F.A., L.Y.F.W. and A.G.S. conceptualised the study, developed the methodology, performed formal analysis, and wrote the main manuscript text. L.Y.F.W. curated the data and prepared all tables and figures. M.P., Y.L., P.R., C.L.W. and A.G.S. validated the results. C.L.W. and A.G.S. provided supervision. All authors reviewed and edited the manuscript.

---

**Supplementary Table 1.** Association between men’s awareness of potential fertility-related adverse health outcomes (Q4) and other characteristics (Q1–3, 6, 9–15 and 19–20).

|  |  | Awareness of potential fertility-related adverse health outcomes (Q4) |  | Group differences |
| --- | --- | --- | --- | --- |
|  |  | Yes | No |  |
| Domain | Characteristics | *n* | *n* | *p*-value |
| 1 | **Age group (Q13)** |  |  | 0.250 |
|  | 18–34 | 1 | 7 |  |
|  | 35–39 | 1 | 9 |  |
|  | 40–44 | 8 | 11 |  |
|  | ≥45 | 5 | 15 |  |
|  | **Highest level of education (Q14)** |  |  | 0.679 |
|  | Finished Secondary school | 0 | 4 |  |
|  | Finished High School or College or equivalent | 3 | 9 |  |
|  | Further qualifications, a University Degree or higher | 12 | 29 |  |
|  | **Ethnicity (Q15)** |  |  | 0.739 |
|  | White (English, Irish, Scottish, etc.) | 12 | 33 |  |
|  | Asian (Indian, Pakistani, Chinese, etc.) | 1 | 3 |  |
|  | Black (Caribbean, African) | 2 | 2 |  |
|  | Mixed (White & Black Caribbean/Asian, etc.) | 0 | 1 |  |
|  | Other ethnic group (Arab, etc.) | 0 | 3 |  |
|  | **UK resident (Q16)** |  |  | **0.018\*** |
|  | Yes | 7 | 32 |  |
|  | No | 5 | 3 |  |
| 2 | **ART status (Q1)** |  |  | 0.408 |
|  | Yes, undergone ART | 7 | 26 |  |
|  | Yes, considering ART | 5 | 8 |  |
|  | No | 9 | 20 |  |
|  | **ART timeframe (Q2)** |  |  | **0.015\*** |
|  | <1 year ago | 7 | 8 |  |
|  | 1–3 years ago | 0 | 9 |  |
|  | 3–5 years ago | 0 | 2 |  |
|  | >5 years ago | 0 | 7 |  |
|  | **ART clinic type (Q3)** |  |  | 0.686 |
|  | NHS clinic | 3 | 9 |  |
|  | Private clinic | 4 | 17 |  |
|  | **Fertility evaluation performed before ART (Q9)** |  |  | 0.056 |
|  | Yes | 3 | 19 |  |
|  | No | 3 | 2 |  |
|  | **Fertility evaluation results discussed (Q10)** |  |  | 1.000 |
|  | Yes | 3 | 16 |  |
|  | No | 0 | 3 |  |
|  | **Fertility evaluation results indicated fertility problems (Q11)** |  |  | 1.000 |
|  | Yes | 3 | 12 |  |
|  | No | 0 | 4 |  |
|  | **Potential adverse health outcomes associated with fertility problems discussed (Q12)** |  |  | 0.081 |
|  | Yes | 2 | 1 |  |
|  | No | 1 | 11 |  |
| 3 | **Concerns about potential fertility-related adverse health outcomes (Q6)** |  |  | 0.122 |
|  | Yes | 15 | 34 |  |
|  | No | 2 | 18 |  |
| 4 | **Concerns about using confidential patient information without consent (Q17)** |  |  | 1.000 |
|  | Yes | 1 | 5 |  |
|  | No | 8 | 23 |  |

\* $p < 0.05$
Abbreviations: ART, assisted reproduction treatment; NHS, National Health Service; Q, question number; UK, United Kingdom

**Supplementary Table 2.**
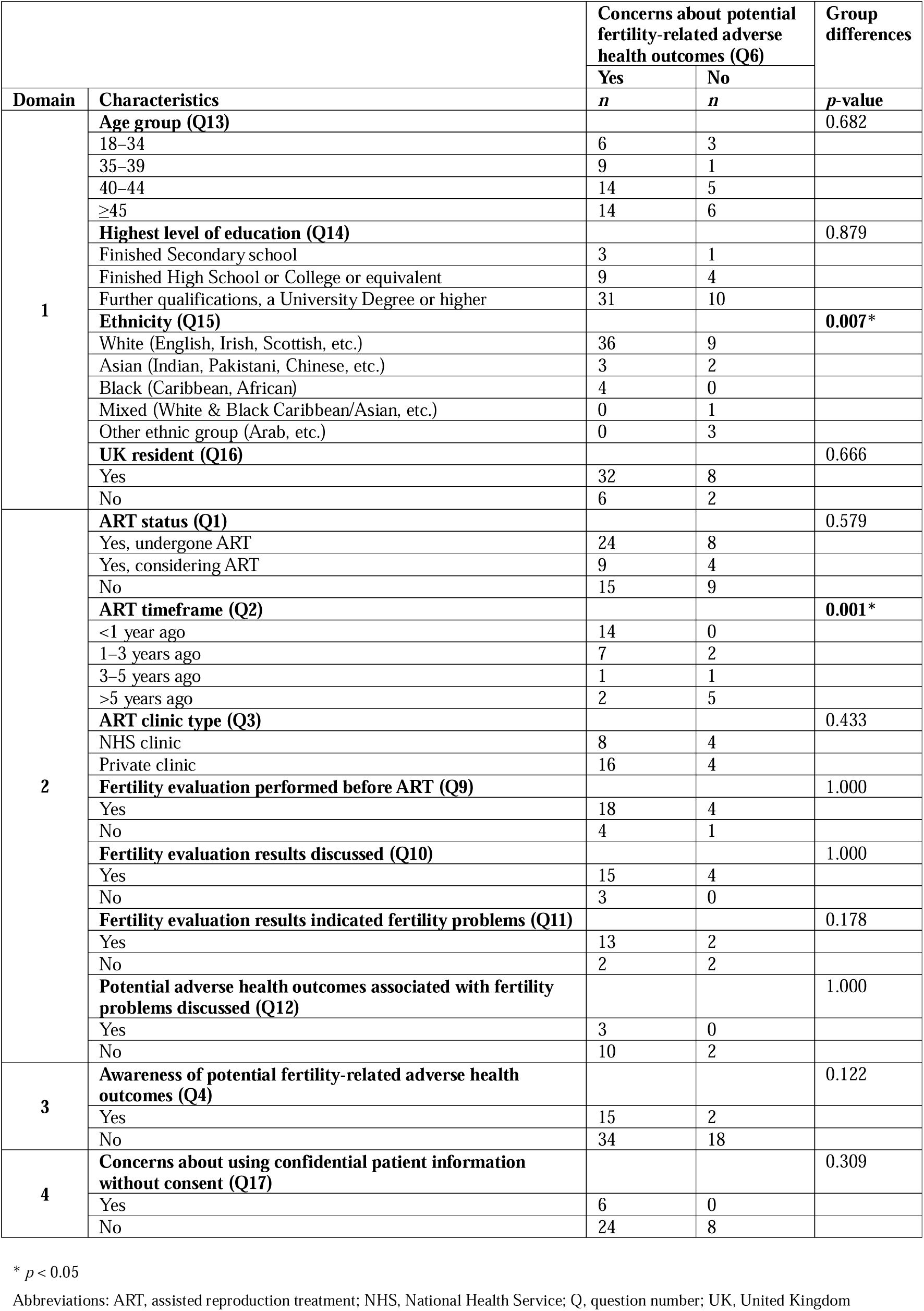
Association between men’s concerns about potential fertility-related adverse health outcomes (Q6) and other characteristics (Q1–4, 9–15 and 19–20).

